# The effect of psychotherapy on the multivariate association between insomnia and depressive symptoms in late-life depression

**DOI:** 10.1101/2025.03.28.25324836

**Authors:** Rafailia Haritos, Vincent Küppers, Fateme Samea, Dieter Riemann, Frank Jessen, Simon B. Eickhoff, Forugh S. Dafsari, Masoud Tahmasian

**Author notes:** **Corresponding Author:** Masoud Tahmasian, Institute of Neuroscience and Medicine, Brain & Behaviour (INM-7), Research Centre Jülich, Wilhelm-Johnen-Straße, 52428 Jülich, Germany. These authors contributed equally (Rafailia Haritos, Vincent Küppers).

## Abstract

**Background:** Late-life depression (LLD) is prevalent in older adults and linked to increased disability, mortality, and suicide risk. Insomnia symptoms are considered common remaining symptoms of LLD following treatment. However, the multivariate relationship between insomnia and depressive symptoms and the impact of psychotherapy on their interrelationship is insufficiently assessed.

**Methods:** We used data from 185 patients with LLD recruited from seven university hospitals in Germany. Participants had undergone eight-week psychotherapy interventions (cognitive behavioral therapy or supportive unspecific intervention). Three regularized canonical correlation analyses (rCCA) assessed the multivariate association between insomnia and depressive symptoms at baseline, post-treatment, and six-month follow-up. rCCA was conducted within a machine learning framework with 100 repeated hold-out splits and permutation tests to ensure robust findings. Canonical loadings and cross-loading difference scores were calculated to examine symptom changes before/after psychotherapy (Holm-Bonferroni corrected p-value < 0.05).

**Results:** At baseline, a moderate association was observed between insomnia and depressive symptoms (r = 0.24). Interestingly, this association slightly increased after the eight-week treatment period (r = 0.42, p_corrected_ = 0.064) and remained significantly elevated at the follow-up session (r = 0.49, p_corrected_ = 0.018). At baseline, anxiety-related depressive symptoms were mainly associated with insomnia, while at post-treatment and follow-up sessions, somatic and negative affective symptoms showed the strongest correlation with insomnia symptoms. Although depressive symptoms significantly improved, insomnia symptoms remained unchanged after psychotherapy.

**Conclusions:** Unlike depressive symptoms, insomnia symptoms did not improve after psychotherapy, highlighting the necessity to target insomnia for effective LLD treatment.

## 1. INTRODUCTION

Late-life depression (LLD) refers to a major depressive disorder (MDD) in individuals aged 60 years or older [1]. It includes a first episode of depression in older adults or recurrent depressive episodes that began earlier in life but re-emerged at an advanced age. LLD is one of the most prevalent mental disorders among older adults, with a 12-month prevalence of approximately 14% [2]. With the rapid growth of the aging population and the high prevalence of LLD, the demand for geriatric mental health care is expected to rise substantially [3]. Beyond this burden, LLD is linked to higher relapse rates and severe health consequences, including accelerated somatic multimorbidity, cognitive decline, and increased mortality [4,5,6,7,8]. Additionally, depressive symptoms vary across the lifespan, posing challenges for interventions. Older adults are more likely to experience somatic and vegetative symptoms, such as insomnia symptoms, including difficulties in sleep initiation, early morning awakening, difficulty staying asleep, and daytime dysfunctions, compared to younger depressed patients [9,10].

Existing evidence suggests that insomnia and depression show a bidirectional relationship. Several meta-analyses and longitudinal studies demonstrated that insomnia symptoms are key risk factors for developing depression [11,12,13]. Conversely, the presence of depression may adversely affect sleep patterns and induce insomnia symptoms [14], which are one of the most prevalent residual symptoms following LLD treatment, with a prevalence of approximately 50% [15,16]. These symptoms have been demonstrated to be associated with the onset, worsening, and recurrence of depressive symptoms, as well as suicidal ideations, particularly in older adults [17,18,19,20]. More severe insomnia symptoms are associated with higher rates of non-response to antidepressant treatment, failed remission, and relapse in LLD [21,22]. To address the association between insomnia and depressive symptoms, multivariate approaches should be employed, as relying solely on the total scores of questionnaires may obscure changes at the individual symptom level of a very heterogeneous clinical condition such as LLD.

Current interventions for LLD include pharmacotherapy and psychotherapy (e.g., Cognitive Behavioral Therapy (CBT)), although suboptimal response to antidepressants and an increased risk of adverse effects, such as hyponatremia, have been reported in older adults [23,24,25]. Thus, the present study aimed to explore the multivariate interrelationship between insomnia and depressive symptoms before and after psychotherapy using canonical correlation analysis (CCA) as conducted previously [26, 27]. This method allows us to assess bidirectional associations between two sets of variables to identify 1) how much insomnia and depressive symptoms are related to each other in patients with LLD at baseline and 2) to determine whether and how psychotherapy treatment may modify their overall multivariate association, as well as each individual variables of both insomnia and depressive scales.

## 2. METHODS

### 2.1. Participants

The current study analyzed a subset from a larger multicenter randomized controlled trial conducted at seven trial sites in Germany (Cognitive Behavioral Therapy for Late-Life Depression - CBTlate trial) [3]. From the original cohort of 251 participants with moderate to severe LLD, we included 185 participants with available scores for the assessment of insomnia and depressive symptoms at 3 time points (baseline, end-of-treatment, and follow-up sessions) (Figure 1a,b). Inclusion criteria were the following: 1) outpatients with age ≥ 60 years; 2) the Geriatric Depression Scale (GDS-30) score > 10; 3) the Quick Inventory of Depressive Symptomatology, Clinician Rating (QIDS-C) score > 10; 4) the Mini-Mental Status Test (MMST) score > 25, to ensure not including patients with severe cognitive impairment; 5) stable pharmacological treatment for at least six weeks before and during the eight weeks of treatment. In view of conditions that could additionally alter sleep patterns, exclusion criteria were: 1) existing comorbidities, including bipolar depression or psychotic disorders; 2) regular use of hypnotics or benzodiazepine medications; 3) other chronic medical/somatic conditions. More details on the in-/exclusion criteria and the interventions in the original trial are available elsewhere [3]. This clinical trial is registered at ClinicalTrials.gov (NCT03735576) and DRKS (DRKS0013769). The study was approved by the Institutional Review Board/Institutional Ethical Committee (IRB/IEC) of the participating sites before the initiation of the trial. All participants provided written informed consent before all study procedures.

**Figure 1.**
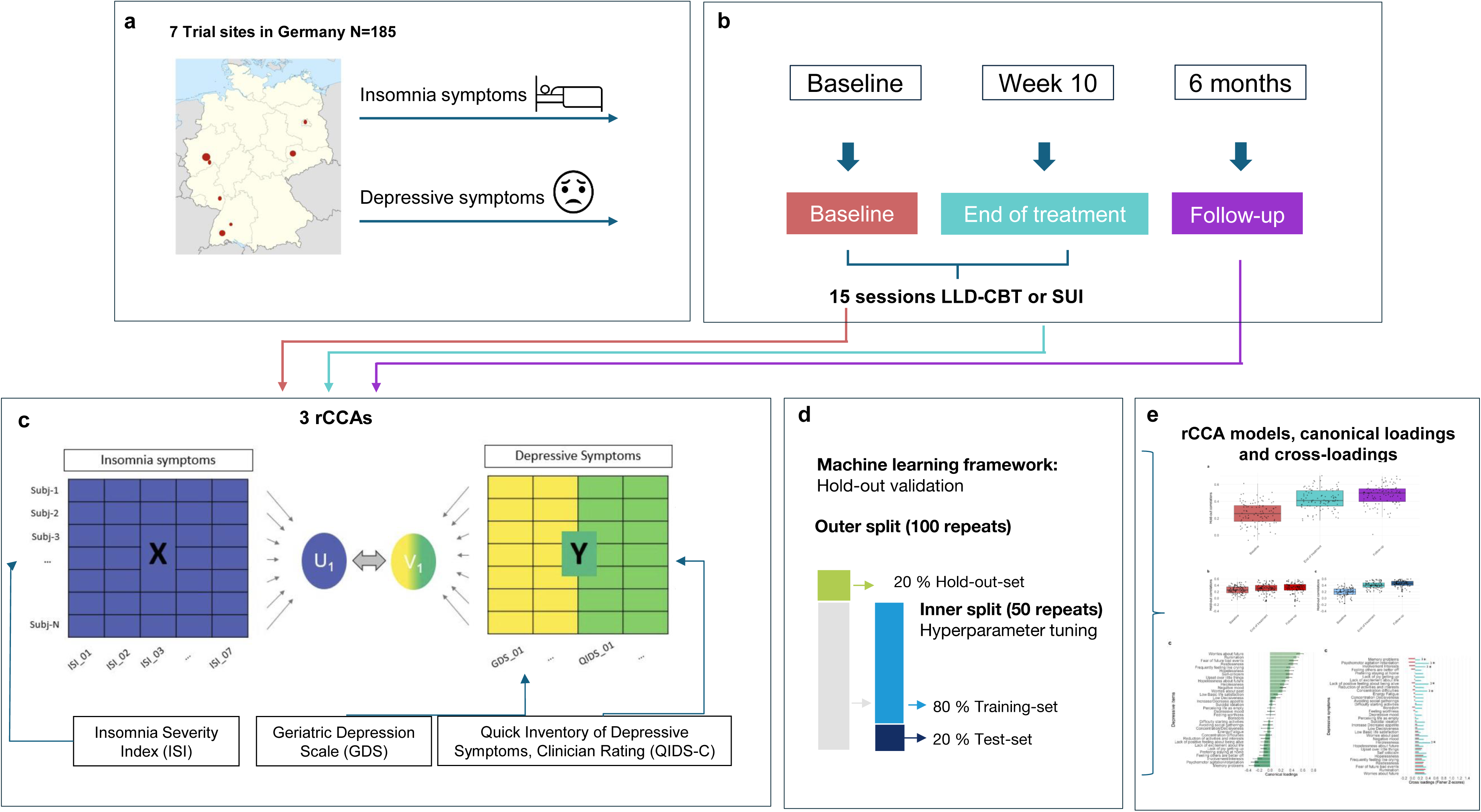
Overview of the study procedure: a) Distribution of participants across trial sites and available data; b) Assessment and intervention time points; c) rCCA model representation of insomnia and depressive symptoms domains; d) Machine learning framework (outer and inner split of data; e) rCCA output: canonical correlation coefficients, canonical loadings and cross-loadings. LLD-CBT: Late-life depression cognitive behavioral therapy; SUI: Supportive unspecific intervention.

### 2.2. Assessments and interventions

Three different assessment time points were considered: baseline scores, at the end of the treatment period (week 10), and the six-month follow-up (Figure 1b).

#### Depressive symptoms

The 30-item Geriatric Depression Scale (GDS) questionnaire [28] was used to assess depressive symptoms. Patients were asked to answer “yes or no” to items indicating how they felt in the past week, including items related to cognitive and affective domains. Scores range from 0 to 30, with higher scores reflecting greater severity of depression. The applied German version has reported a high internal consistency (Cronbach’s alpha = 0.91) and a high test-retest validity coefficient of 0.83 [29].

The QIDS-C [30] was used as a secondary measure of depressive symptoms, providing further information on the somatic domain of depressive symptoms. It is a 16-item clinician rating scale that assesses DSM-IV criterion diagnostic symptoms of MDD using a Likert-type scale ranging from 0 (none) to 3 (very much). The total score ranges from 0 to 27, with higher scores indicating greater depression severity. Single-item responses are converged into scores in nine criterion domains, including sad mood, concentration, self-criticism, suicidal ideation, interest, energy/fatigue, sleep disturbance (initial, middle, and late insomnia or hypersomnia), decrease/increase in appetite/weight and psychomotor agitation or retardation. An acceptable internal consistency (Cronbach’s alpha = 0.77) has been reported for the applied German version [31].

#### Insomnia symptoms

Insomnia symptoms were measured using the Insomnia Severity Index (ISI) [32]. This seven-item self-report questionnaire assesses difficulty falling asleep, staying asleep, or waking up too early, overall satisfaction with the sleep pattern, interference of sleep problems with daily functioning, how noticeable sleep problems are to others, and worry/distress caused by the sleep problems. A Likert-type scale is used, with scores ranging from 0 (none) to 4 (very much), and higher scores indicating greater severity of insomnia symptoms. Total scores range from 0 to 28, with higher scores reflecting higher symptom severity. The applied German version has been shown to have a high internal consistency (Cronbach’s alpha = 0.81) [33].

#### Psychotherapeutic treatment

Scores on the GDS and ISI were considered at the baseline assessment before treatment and after receiving either the LLD-specific cognitive behavioral therapy (LLD-CBT) or the supportive unspecific intervention (SUI). Both groups underwent a structured manual-based treatment consisting of 15 sessions, conducted twice weekly over an eight-week period. Further details on the interventions have been published elsewhere [3,34].

### 2.3. Multivariate and statistical analyses

#### Regularised canonical correlation analyses (rCCA)

We applied CCA and conducted three separate analyses for the baseline, end-of-treatment, and follow-up individual scores of insomnia and depressive symptoms. CCA is a multivariate, data-driven statistical method integrating various symptoms into a single model to identify latent dimensions where specific items from both sets are maximally correlated [35]. A “canonical variate” represents the specific linear combination of variables within each set, and a pair of canonical variates constitutes a mode. The “canonical loading” scores were computed to interpret the modes of the CCA models, corresponding to Pearson correlations between each input variable and the canonical variates (Figure 1c).

One set of variables (domain X) represented the insomnia dimension, which consisted of scores from seven items of ISI. The other set of variables (domain Y) represented depressive symptoms, comprising scores from 26 GDS items and eight QIDS-C items. Four items of the GDS (Sadness, feeling downhearted, feeling without energy, not feeling clear-minded) were excluded, as they showed an extremely skewed distribution in our sample. Notably, the sleep disturbance criterion domain (which includes initial, middle, and late insomnia or hypersomnia) from the QIDS-C was excluded, as performed previously [36]. This decision was made to avoid including any sleep-related items within the depressive domain, as doing so could result in an inaccurately higher association between the sets. Since both LLD-CBT and SUI significantly reduced depressive symptoms without evidence for the superiority of LLD-CBT in the initial trial [3,35], all participants were pooled for the current analyses, regardless of the allocation to the treatment groups. The analyses were implemented by using the CCA/PLS Toolkit (v1.0.0, https://github.com/anaston/cca_pls_toolkit/) in MATLAB Runtime (v9.14 (R2023b)) [37]. Age, gender, and trial site were included in the analysis as confounding variables.

To address potential limitations of CCA (i.e., overfitting and multicollinearity among variables in each behavioral domain), a regularized version of CCA (rCCA) was implemented within a machine learning framework [26,27]. A multiple hold-out framework was utilized by dividing the dataset into an 80% optimization set and a 20% hold-out set, using 100 random repetitions. This approach evaluates the model’s generalizability by examining how well the associations identified in the training set can apply to an independent test set. In this process, the hold-out set was projected onto the optimal rCCA model obtained from the optimisation set, aiming to estimate the canonical correlation of the hold-out set. The optimisation set was further split into an 80% training set and a 20% test set through an inner split, repeated 50 times. During this inner split, optimal hyperparameters were estimated using L2 norm regularisation (Figure 1d). This additional constraint forces the weights to be small but not zero. By mitigating multicollinearity, these hyperparameters help enhance the stability of the rCCA weights [38].

We further assessed the longitudinal stability and alteration of the associations across time points by testing whether pre-treatment associations persist post-treatment and whether new relationships emerge after treatment. Therefore, the end-of-treatment and follow-up data were projected onto the weights derived from the baseline rCCA model. Additionally, the baseline and follow-up data were projected onto the weights of the end-of-treatment model. The model was initially trained at the baseline, after which it was tested on the hold-out set from the baseline time point. Subsequently, the model was tested on the hold-out sets from the other two time points, and this process was repeated for the end-of-treatment model. This process was repeated for all 100 hold-out repetitions to ensure the robustness of rCCA correlation results.

#### Fisher’s Z transformation and statistical comparisons

We used Fisher’s Z transformation to compare the correlation coefficients from our rCCA analyses before and after treatment, assessing whether the treatment significantly altered the association between insomnia and depressive symptoms. First, we normalized canonical correlation coefficients, loadings, and cross-loadings by computing their Fisher’s Z scores. Using Fisher’s Z-test, we then compared canonical correlation coefficients from baseline to end of treatment and follow-up, applying Holm-Bonferroni correction for multiple comparisons. To further identify symptom-specific changes, we examined shifts in canonical loadings (reflecting how individual symptoms contribute to their domains) and cross-loadings (indicating how symptoms of one domain relate to the overall construct of the other). Delta scores (end of treatment – baseline Fisher’s Z) were tested using Fisher’s Z-test to detect significant changes, highlighting symptom-level interactions influenced by treatment.

## 3. RESULTS

### 3.1 Descriptive statistics

The data set comprised 185 participants, with an average age of 69.7 years old (SD=7.1) of whom 66% were female. Patients were recruited from seven university hospitals in Germany (Table 1).

**Table 1.**
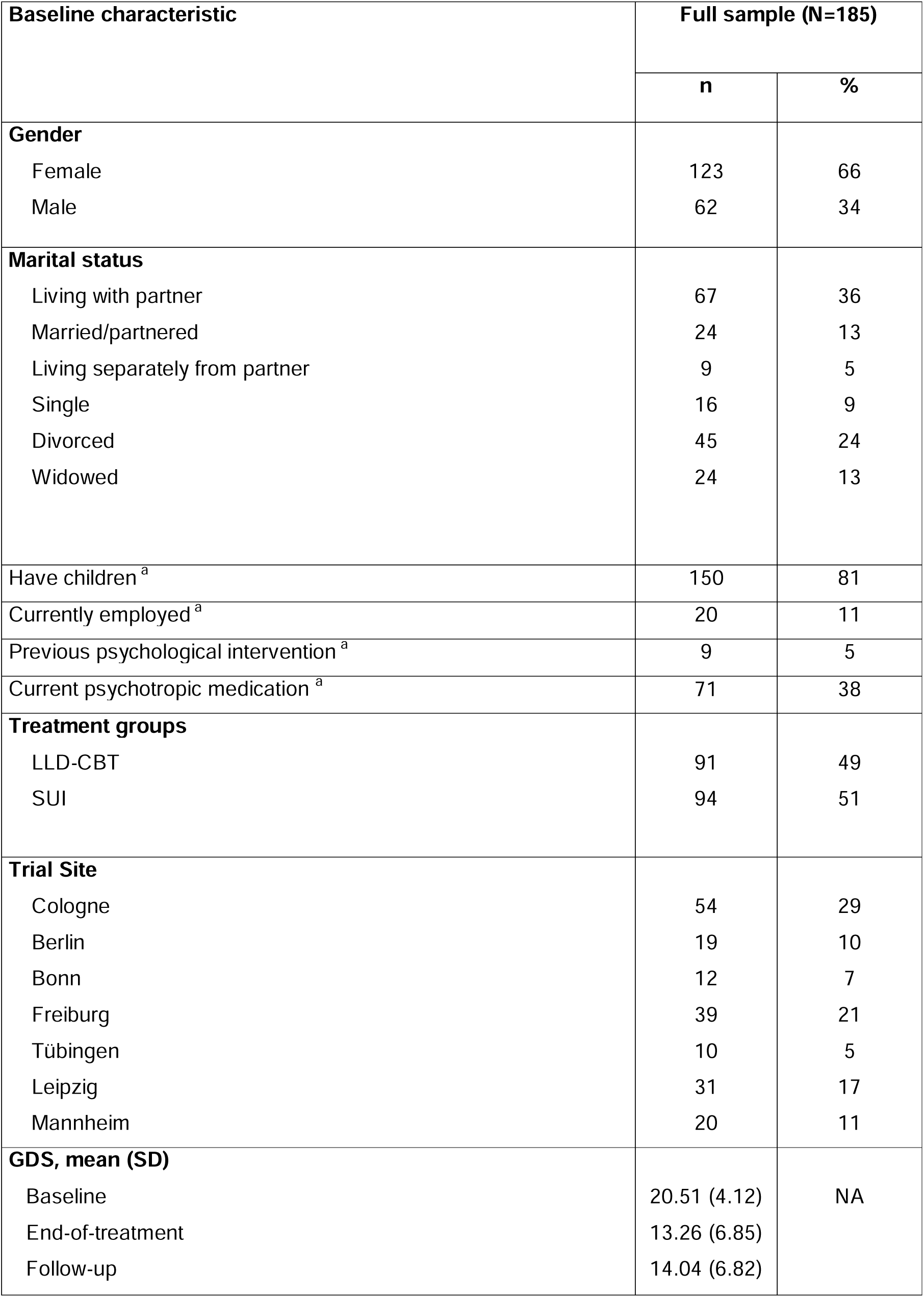
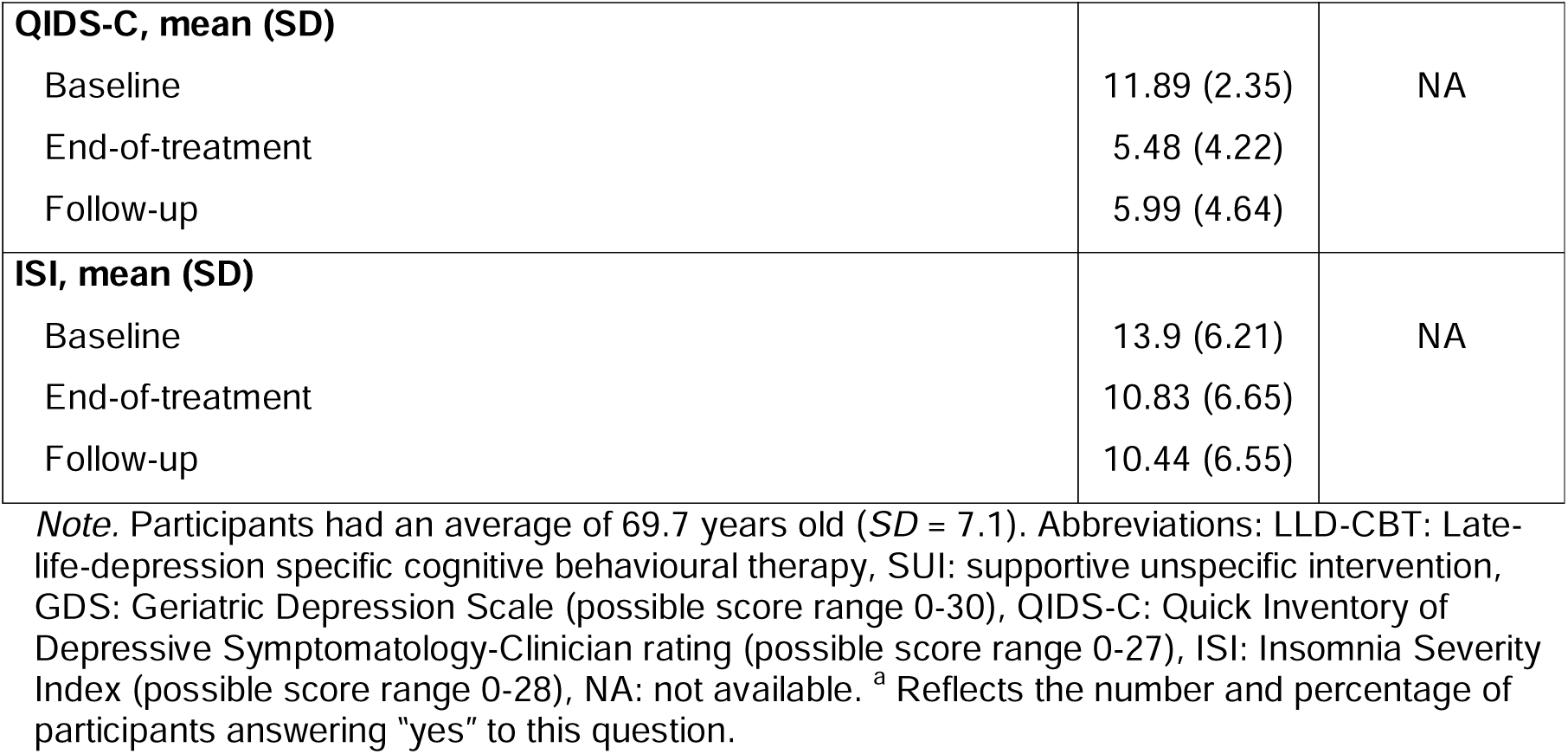
Patient characteristics.

| Baseline characteristic | Full sample (N=185) |  |
| --- | --- | --- |
|  | n | % |
| **Gender** |  |  |
| Female | 123 | 66 |
| Male | 62 | 34 |
| **Marital status** |  |  |
| Living with partner | 67 | 36 |
| Married/partnered | 24 | 13 |
| Living separately from partner | 9 | 5 |
| Single | 16 | 9 |
| Divorced | 45 | 24 |
| Widowed | 24 | 13 |
| Have children <sup>a</sup> | 150 | 81 |
| Currently employed <sup>a</sup> | 20 | 11 |
| Previous psychological intervention <sup>a</sup> | 9 | 5 |
| Current psychotropic medication <sup>a</sup> | 71 | 38 |
| **Treatment groups** |  |  |
| LLD-CBT | 91 | 49 |
| SUI | 94 | 51 |
| **Trial Site** |  |  |
| Cologne | 54 | 29 |
| Berlin | 19 | 10 |
| Bonn | 12 | 7 |
| Freiburg | 39 | 21 |
| Tübingen | 10 | 5 |
| Leipzig | 31 | 17 |
| Mannheim | 20 | 11 |
| **GDS, mean (SD)** |  |  |
| Baseline | 20.51 (4.12) | NA |
| End-of-treatment | 13.26 (6.85) |  |
| Follow-up | 14.04 (6.82) |  |
| **QIDS-C, mean (SD)** |  |  |
| Baseline | 11.89 (2.35) | NA |
| End-of-treatment | 5.48 (4.22) |  |
| Follow-up | 5.99 (4.64) |  |
| **ISI, mean (SD)** |  |  |
| Baseline | 13.9 (6.21) | NA |
| End-of-treatment | 10.83 (6.65) |  |
| Follow-up | 10.44 (6.55) |  |

*Note.* Participants had an average of 69.7 years old ( $SD = 7.1$ ). Abbreviations: LLD-CBT: Late-life-depression specific cognitive behavioural therapy, SUI: supportive unspecific intervention, GDS: Geriatric Depression Scale (possible score range 0-30), QIDS-C: Quick Inventory of Depressive Symptomatology-Clinician rating (possible score range 0-27), ISI: Insomnia Severity Index (possible score range 0-28), NA: not available. <sup>a</sup> Reflects the number and percentage of participants answering “yes” to this question.

### 3.2 Alteration of multivariate association between insomnia and depressive symptoms after psychotherapy

At baseline, a hold-out set correlation of r = 0.24 (SD = 0.13) was observed between insomnia symptoms and depressive symptoms. By the end of the treatment, this correlation increased slightly (r = 0.42, SD = 0.12) and remained consistently high at the six-month follow-up session (r = 0.48, SD = 0.12) (Figure 2a). Notably, the increase in the hold-out correlation from baseline to the end of treatment was not significant after applying the Holm-Bonferroni correction (p_corrected_ = 0.064). However, the increase from baseline to follow-up was significant (p_corrected_ = 0.018). The projection of the baseline model to the post-treatment model shows a similar pattern, reflecting the stability of the identified associations (Figure 2b). Conversely, when projecting the end of the treatment model to the baseline hold-out set, a decrease in the correlation can be observed, indicating new association patterns that were not present at baseline (Figure 2c).

**Figure 2.**
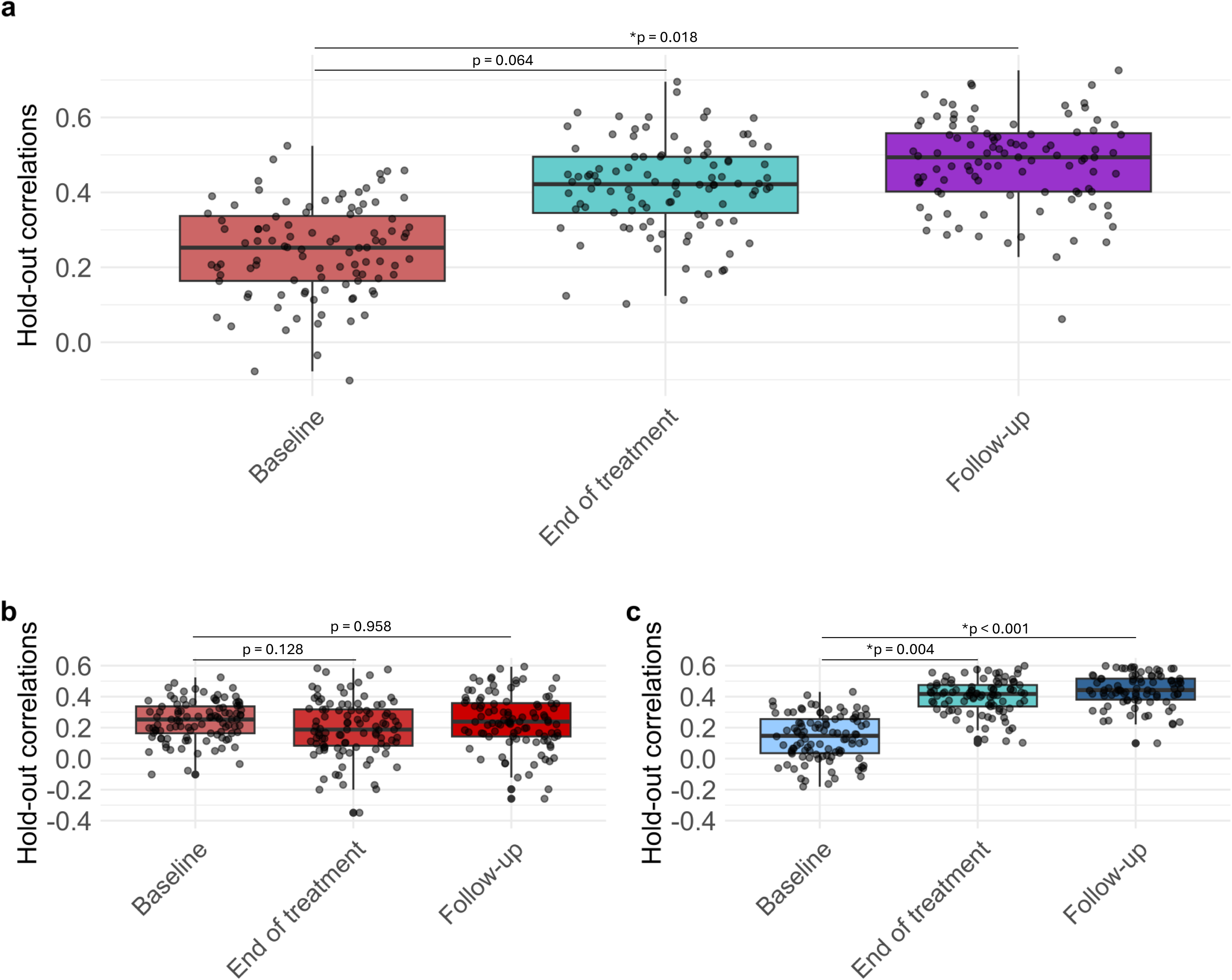
Hold-out correlations of each regularised canonical correlation analysis (rCCA) model: a) rCCA model between insomnia and depressive symptoms for baseline, end-of treatment and follow-up time points; b) Testing the baseline rCCA model on each time point’s hold-out set; c) Testing the end-of-treatment model on each time point’s hold-out set. Dots represent the hold-out correlations for each of the 100 repetitions. Black horizontal lines indicate the mean hold-out correlation coefficients. (*) Holm-Bonferroni corrected significant differences (p-value < 0.05).

At baseline, insomnia symptoms were positively associated with depressive symptoms, indicating that greater insomnia symptom severity was associated with more severe depressive symptoms. Higher canonical loadings were observed for general dissatisfaction with the sleep pattern (r = 0.88), the interference of sleep problems with daily functioning (r = 0.81), and difficulty staying asleep (r = 0.81) (Figure 3a-b). Higher canonical loadings were observed mainly for anxiety-related depressive symptoms, such as worrying about the future (r = 0.52), excessive rumination (r = 0.44), fear of the occurrence of future bad events (r = 0.42) or general feelings of restlessness (r = 0.41). Decreased involvement and interest in daily life (r = −0.31), increased hopelessness about the future (r = −0.29), negative mood (r = −0.25), psychomotor agitation or retardation (r = −0.25), or increased memory problems (r = −0.22) were strongly associated with less insomnia symptoms (Figure 3c-d).

**Figure 3.**
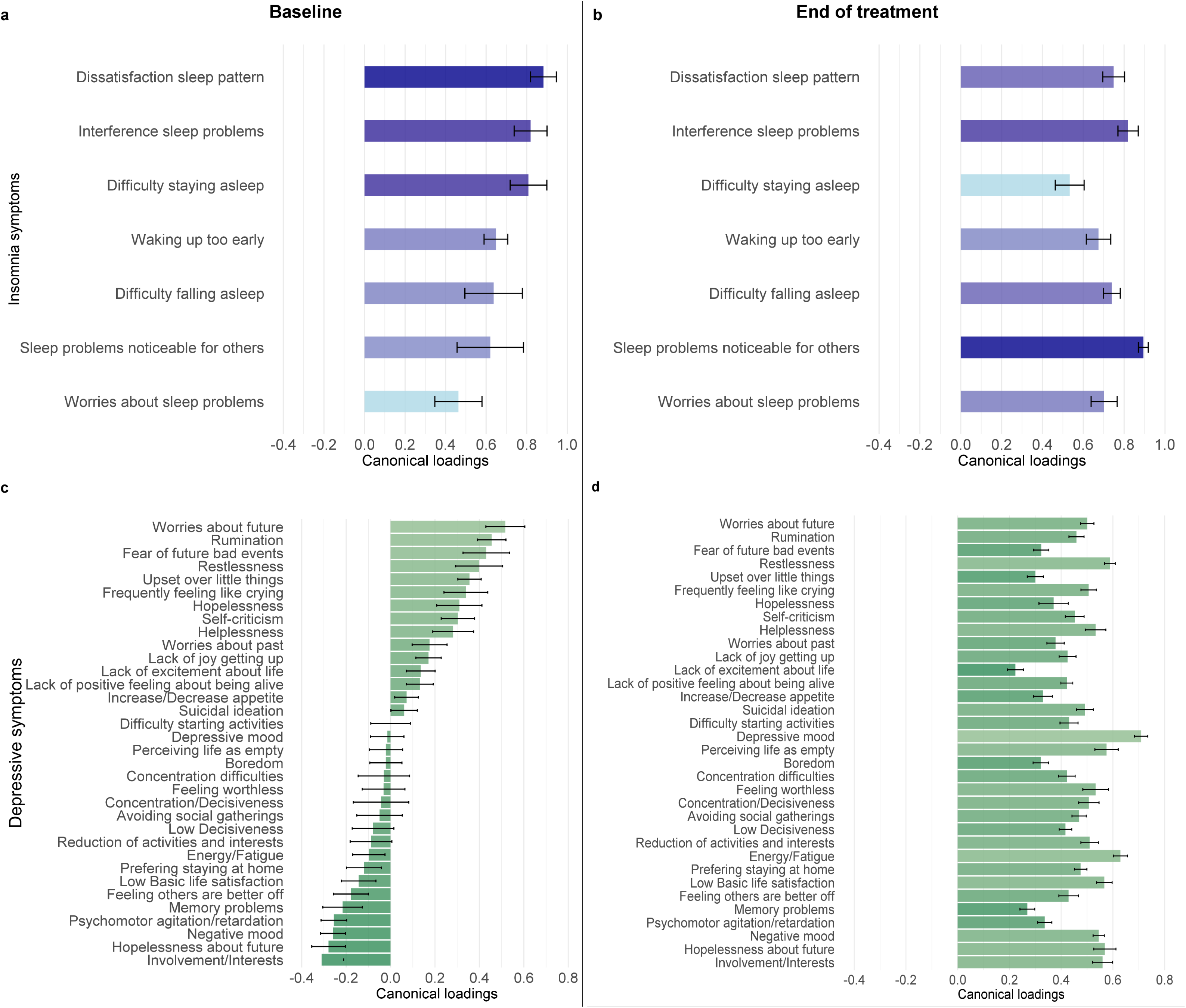
Canonical loadings before and after psychotherapeutic treatment: a) Canonical loadings of insomnia symptoms at baseline; b) Canonical loadings of insomnia symptoms at the end of treatment; c) Canonical loadings of depressive symptoms at baseline; d) Canonical loadings of depressive symptoms at the end of treatment. Canonical loadings correspond to Pearson correlations between the input variables and the canonical variates. Negative loadings of depressive symptoms indicate more insomnia symptoms, and less depressive symptoms, while positive loadings indicate more insomnia symptoms and more depressive symptoms. Black lines represent error bars (one standard deviation below or above the mean).

At the end of treatment, the canonical loadings for all insomnia symptoms were positively associated with depressive symptoms. The strongest associations were observed for sleep problems being noticeable to others (r = 0.89), sleep problems interfering with daily functioning (r = 0.82), general dissatisfaction with sleep pattern (r = 0.75), and difficulty falling asleep (r = 0.74) (Figure 3a-b). All depressive symptoms following psychotherapy intervention showed a positive association with insomnia symptoms. The highest loadings were seen in negative affective symptoms, such as overall depressive mood (r = 0.71), restlessness (r = 0.59), perceiving life as empty (r = 0.58), feeling hopeless about the future (r = 0.57) and having a low basic life satisfaction (r = 0.57). Additionally, somatic symptoms, such as low energy and high fatigue, were also strongly and positively associated with insomnia symptoms (r = 0.63) (Figure 3c-d).

At follow-up, the loadings for insomnia symptoms remained consistent with the canonical loadings at the end of treatment. Similarly, all depressive symptoms at follow-up remained positively associated with insomnia symptoms (Supplementary Material).

### 3.3 Changes in canonical loadings and cross-loadings after psychotherapy

The canonical loadings of most depressive items changed significantly after psychotherapy compared to baseline (corrected p-values between 0.002 and 0.005) (Figure 4a). The calculated difference (delta) scores for the “canonical loadings” revealed the most significant changes in items related to reduced activities (delta = 0.501, p < 0.001), interests, and involvement in daily life (delta = 0.546, p < 0.001), psychomotor agitation or retardation (delta = 0.54, p < 0.001), memory problems (delta = 0.543, p < 0.001), difficulty concentrating and decisiveness (delta = 0.504, p < 0.001). In contrast, the canonical loadings of insomnia symptoms showed no significant changes after treatment for any items (Figure 4b).

**Figure 4.**
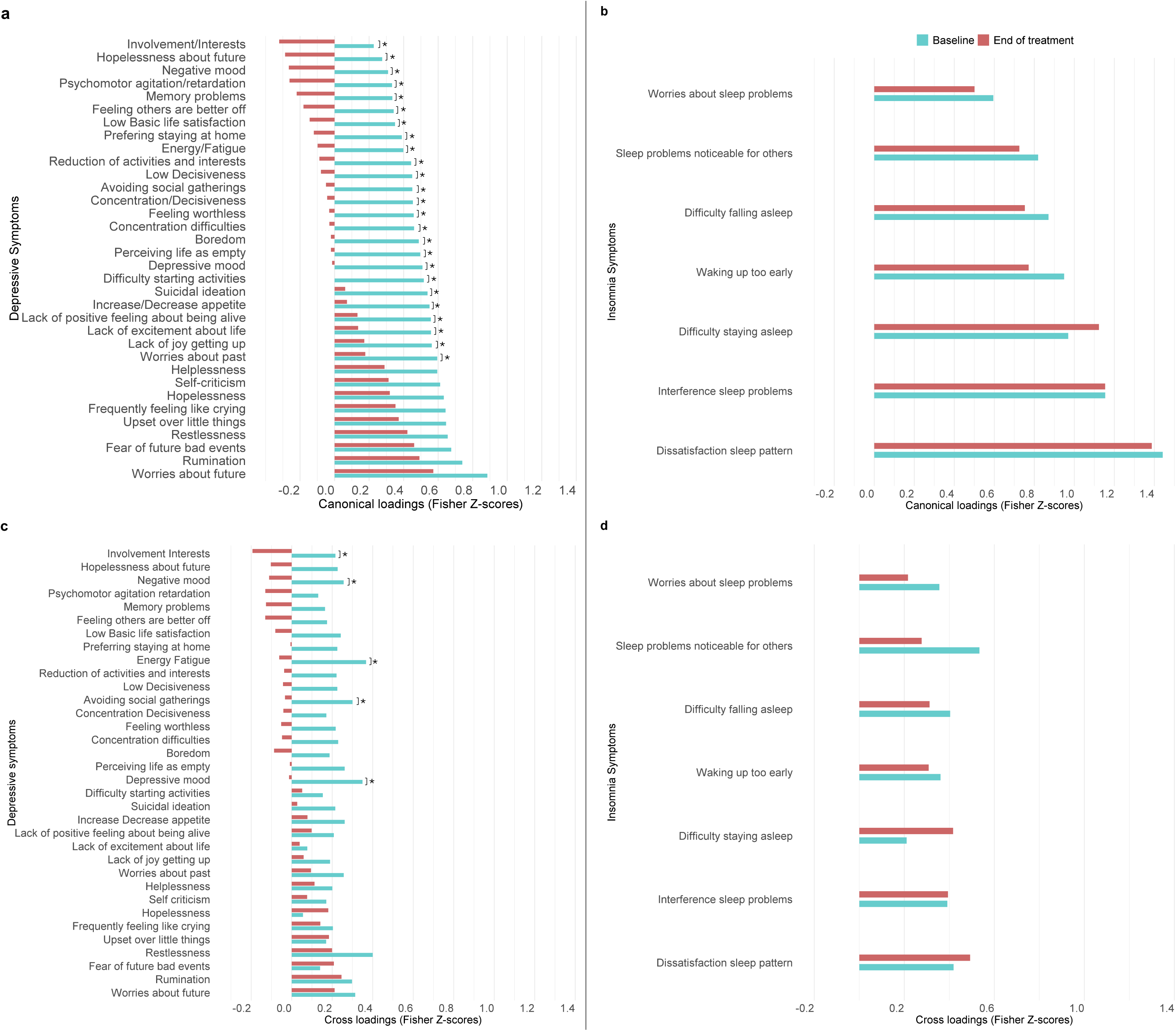
Changes in canonical loadings and cross-loadings before and after psychotherapeutic treatment: a) Fisher z-scores of canonical loadings at baseline and after treatment for depressive symptoms; b) Fisher z-scores of canonical loadings at baseline and after treatment for insomnia symptoms; c) Fisher z-scores of cross-loadings at baseline and after treatment for depressive symptoms; d) Fisher z-scores of cross-loadings at baseline and after treatment for insomnia symptoms. (*) Holm-Bonferroni corrected significant differences between time points (p-value < 0.05).

With regard to the cross-loadings of depressive symptoms (as the association with the insomnia canonical variate), significant changes after psychotherapy were observed for psychomotor agitation or retardation (delta = 0.516, p < 0.001), decrease in involvement and interests (delta = 0.415, p < 0.001), concentration difficulties (delta = 0.371, p = 0.001), feelings of helplessness (delta = 0.298, p = 0.007), and memory problems (delta = 0.294, p = 0.008) (Figure 4c). Conversely, the cross-loadings of insomnia symptoms (as the association with the depressive symptoms canonical variate) did not show any significant changes (Figure 4d), which emphasized that the insomnia symptoms pattern remained stable and independent from changes in depressive symptoms following CBT/SUI psychotherapy.

## 4. DISCUSSION

We found a moderate association between insomnia and depressive symptoms at baseline, providing further evidence of their bidirectional relationship in LLD. This association slightly increased at the end of treatment and the six-month follow-up. While we observed improvement in depressive symptoms, insomnia symptoms did not change after psychotherapy.

Existing evidence suggested several shared underlying mechanisms for the insomnia-depressive symptoms link, including shared genetic factors, hyperactivity of the arousal system, disruptions in synaptic plasticity, alterations in the biological timekeeping system, and shared macroscale neuroanatomical abnormalities between insomnia and depression [39,40,41,42,43]. We observed a trend of increasing multivariate associations between insomnia and depressive symptoms after psychotherapy. Additionally, by projecting the end of the treatment model to the baseline test set, we observed a decrease in the correlation, highlighting increased or new patterns of association after psychotherapy, which could not be captured by the baseline model. Since the interventions primarily addressed depressive symptoms rather than insomnia, unresolved sleep problems could have exacerbated specific depressive symptoms, thereby strengthening their interrelationship. Changes between the baseline and the end of treatment model were not significant after correction, probably due to our small sample size and high data variability. It has been shown that antidepressant treatment and commonly used psychotherapy interventions alleviate certain types of depressive symptoms but fail to fully resolve insomnia symptoms in depressed patients [44,45]. These residual insomnia symptoms increase vulnerability to depressive relapse or recurrence and a lower likelihood for patients to have full benefit from treatment [46,47]. Residual insomnia symptoms are then often treated with hypnotic medications or particular antidepressants despite the risks of side effects (e.g., daytime impairment, amnesia, falling down, and dependency), particularly in older adults, who are nearly five times more likely to receive prescriptions of medications than younger individuals [48,49,50]. Given these concerns, a non-pharmacological approach, such as cognitive behavioral therapy for insomnia (CBT-I)—which includes cognitive therapy, stimulus control, sleep restriction, sleep hygiene, and relaxation techniques—may be an additional and effective first-line treatment and could be integrated into clinical practice for older adults with LLD to prevent residual insomnia [16,51]. CBT-I is suggested as an early effective intervention in mood disorders [52]. The stable association observed between insomnia and depressive symptoms after a six-month follow-up suggests a persistent relationship and symptom interaction pattern. Interestingly, a study based on the Growth mixture modeling demonstrated that characteristics of insomnia can predict long-term depression outcomes. In particular, patients with lower insomnia severity were the optimal responders to psychotherapy and exhibited more favorable treatment responses, even at a two-year follow-up [53].

At baseline, our canonical loadings indicated a positive relationship between insomnia symptoms and anxiety-related depressive symptoms, particularly excessive worry, rumination, and feelings of restlessness (Figure 4a). Excessive uncontrollable worry, rumination, emotion dysregulation, and intrusive thoughts have been associated with increased insomnia symptoms in previous studies due to hyperarousal conditions [54,55,56,57], also for older patients [58]. In this study, after psychotherapy and at follow-up, this association shifted, with insomnia symptoms becoming more strongly linked to negative affective symptoms, such as low mood, negative outlook, and reduced life satisfaction, as well as somatic symptoms, including low energy and heightened fatigue (Figure 4a-b). This further suggests that while CBT/SUI treatment may have focused more on cognitive aspects, sleep-related and somatic symptoms remained unchanged, contributing to persistent negative mood. Zhao and colleagues (2021) [59] found that older patients with insomnia were more likely to experience somatic symptoms, such as low energy, which in turn were linked to persistent negative affect.

Individuals with the same severity of depression can exhibit substantial differences in how their symptoms interact [60]. A novel “person-specific symptom network” model suggests that examining individual symptom relationships could help identify maladaptive patterns, enabling personalized treatment approaches to weaken these persistent associations [61]. We observed that psychomotor agitation or retardation, memory and concentration difficulties, and decreased engagement in daily activities were negatively associated with insomnia at baseline but became positively linked by the end of treatment and follow-up. These symptoms showed the highest cross-loading delta scores, reflecting significant shifts in their relationship with insomnia over time (Figure 4c-d). Psychomotor agitation has been linked to depression in older adults, often alongside increased insomnia symptoms [62]. Agitated depression may represent a distinct LLD subtype, where heightened agitation initially masks perceived sleep difficulties, explaining the negative correlation at baseline. After therapy, patients may have gained a greater awareness of their sleep disturbances, leading to the observed change in the association. Current models of insomnia emphasize its role in hyperarousal driven by central and peripheral activation of the stress system and its involvement in GABAergic transmission [63]. This dysregulation may contribute to symptoms such as psychomotor agitation or retardation.

Recognizing our limitations can help guide future research to strengthen and expand upon our findings. For example, recall/response bias is common in self-report questionnaires. Moreover, psychotherapy may enhance introspection and self-awareness, enabling patients to better recognize the connection between sleep disturbances and depressive symptoms, making this relationship more apparent after the treatment [64]. Additionally, while individual depressive symptoms showed varying canonical loadings, all depressive symptoms exhibited high loadings post-treatment and at follow-up. Notably, individuals with mood disorders often report excessive sleep disturbances and poor sleep quality, though objective measures like polysomnography may show different results [65,66]. This study relied solely on the ISI, which focuses on insomnia; incorporating additional subjective and objective sleep measures, such as sleep duration and efficiency, could provide a more comprehensive assessment on the sleep-depression relationship. Future research should also compare different treatment modalities, particularly LLD-CBT, versus insomnia-specific interventions (e.g., CBT-I). Neuroimaging data could further clarify the intervention’s impact and reveal underlying neurobiological substrates. Patients in this study received either LLD-CBT or SUI, but separate rCCA models for each group were not statistically valid due to sample size limitations. Larger datasets are needed to confirm these findings and perform clustering approaches based on the link between sleep and depression [67,68].

In conclusion, this study demonstrated increased multivariate association between insomnia and depressive symptoms in LLD patients following psychotherapy, alongside a shift in the pattern of depressive symptoms, while insomnia symptoms remained unchanged. These findings emphasize the need to consider their relationship when assessing treatment effects. Furthermore, the findings underscore the value of multivariate approaches in examining symptom interactions, ultimately aiming to enhance clinical treatment.

## Data Availability

Due to ethics restrictions, the data supporting this study’s findings are not publicly available, but they are available from Dr. Forugh Salimi Dafsari upon reasonable request with individual permission from the local institutions’ ethics board.

## Acknowledgments

The authors would like to thank all participating patients, therapists, and outcome evaluators who participated in the study.

## Author Contribution

Conception and study design: R.H., V.K., F.D., and M.T; Preprocessing and data analysis: R.H., V.K., F.S.; Interpretation F.S., F.D., D.R., S.B.E., F.J., and M.T. Paper writing and editing: all authors.

