## Supplemental file for "The effect of psychotherapy on the multivariate association between insomnia and depressive symptoms in late-life depression"

Rafailia Haritos et al. 2025

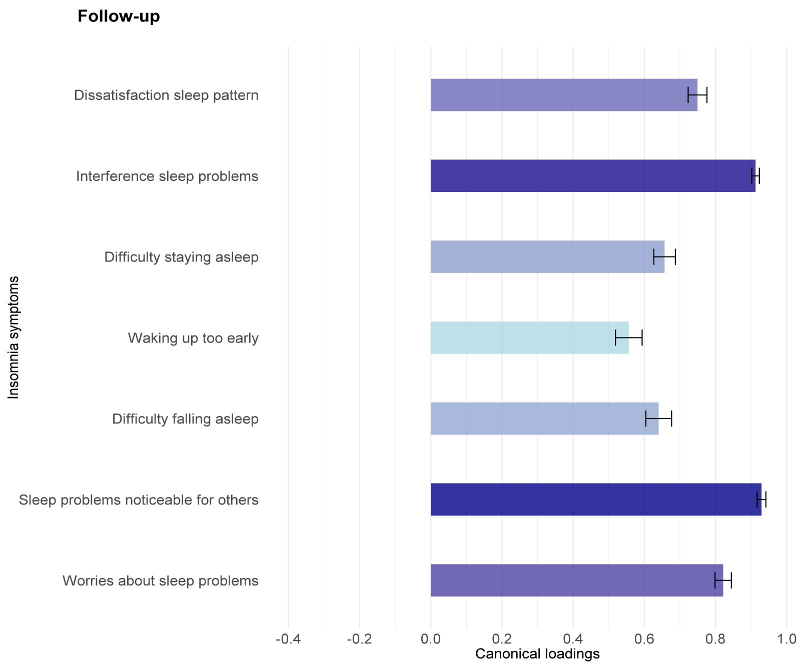


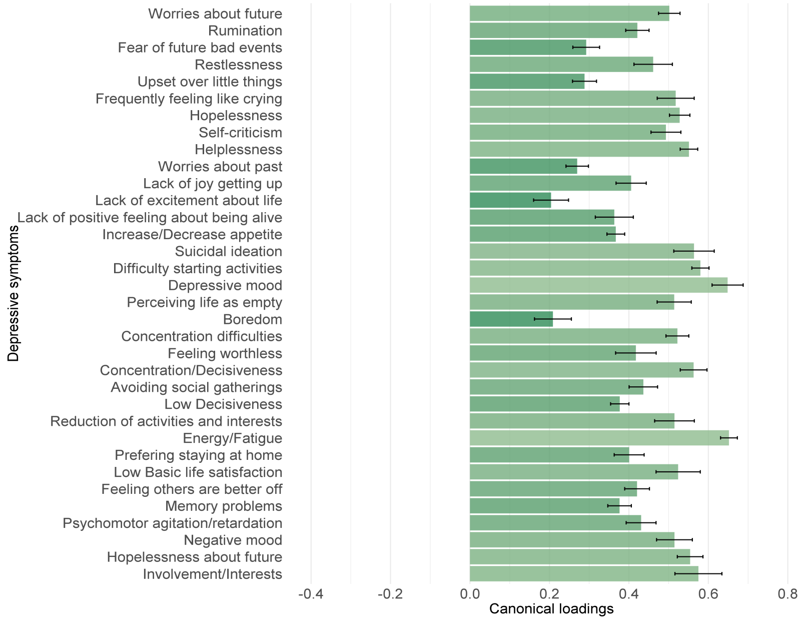


**Figure S1. Canonical loadings at Follow-up.** Canonical loadings of insomnia symptoms and depressive symptoms at follow-up. Canonical loadings correspond to Pearson correlations between the input variables and the canonical variates. Positive loadings indicate more insomnia symptoms and more depressive symptoms. Black lines represent error bars (one standard deviation below or above the mean).
